# Accurate Prediction of COVID-19 using Chest X-Ray Images through Deep Feature Learning model with SMOTE and Machine Learning Classifiers

**DOI:** 10.1101/2020.04.13.20063461

**Authors:** Rahul Kumar, Ridhi Arora, Vipul Bansal, Vinodh J Sahayasheela, Himanshu Buckchash, Javed Imran, Narayanan Narayanan, Ganesh N Pandian, Balasubramanian Raman

## Abstract

According to the World Health Organization (WHO), the coronavirus (COVID-19) pandemic is putting even the best healthcare systems across the world under tremendous pressure. The early detection of this type of virus will help in relieving the pressure of the healthcare systems. Chest X-rays has been playing a crucial role in the diagnosis of diseases like Pneumonia. As COVID-19 is a type of influenza, it is possible to diagnose using this imaging technique. With rapid development in the area of Machine Learning (ML) and Deep learning, there had been intelligent systems to classify between Pneumonia and Normal patients. This paper proposes the machine learning-based classification of the extracted deep feature using ResNet152 with COVID-19 and Pneumonia patients on chest X-ray images. SMOTE is used for balancing the imbalanced data points of COVID-19 and Normal patients. This non-invasive and early prediction of novel coronavirus (COVID-19) by analyzing chest X-rays can further be used to predict the spread of the virus in asymptomatic patients. The model is achieving an accuracy of 0.973 on Random Forest and 0.977 using XGBoost predictive classifiers. The establishment of such an approach will be useful to predict the outbreak early, which in turn can aid to control it effectively.

## Introduction

COVID-19, commonly referred to as coronavirus disease-19 has emerged as a fatal SARS infection^1^ over the past few months. With the place of origin being identified as Wuhan, China, the disease now stands as being declared ‘pandemic’ by the World Health Organization (WHO)^2–4^. The symptoms of the disease resemble a typical viral, respiratory infection with an incubation time of214days. However, as this disease progresses, the affected experiences shortness of breath, nausea culminating in pneumonia and multiple organ failure^3, 5^.

Given the deadly and debilitating condition that it is, the entire world now is at high alert with thousands of cases being identified each day. Going by WHO statistics, as of now, there have been about 1.84 million identified cases with over 1, 13, 329 deaths^6^. Country-wise (top 10 India) statistical COVID-19 cases as of April 12, 2020, presented in Table 1^7^. According to WHO^8^, geographical statistics of the confirmed COVID-19 cases have registered as of April 12, 2020 as depicted in the Figure 1. International travel history and close contact with the infected have been identified as the causes of the worldwide spread of the disease. An enormous amount of effort is being put into developing vaccines and drugs to help treat the infection^9, 10^.

**Table 1.**
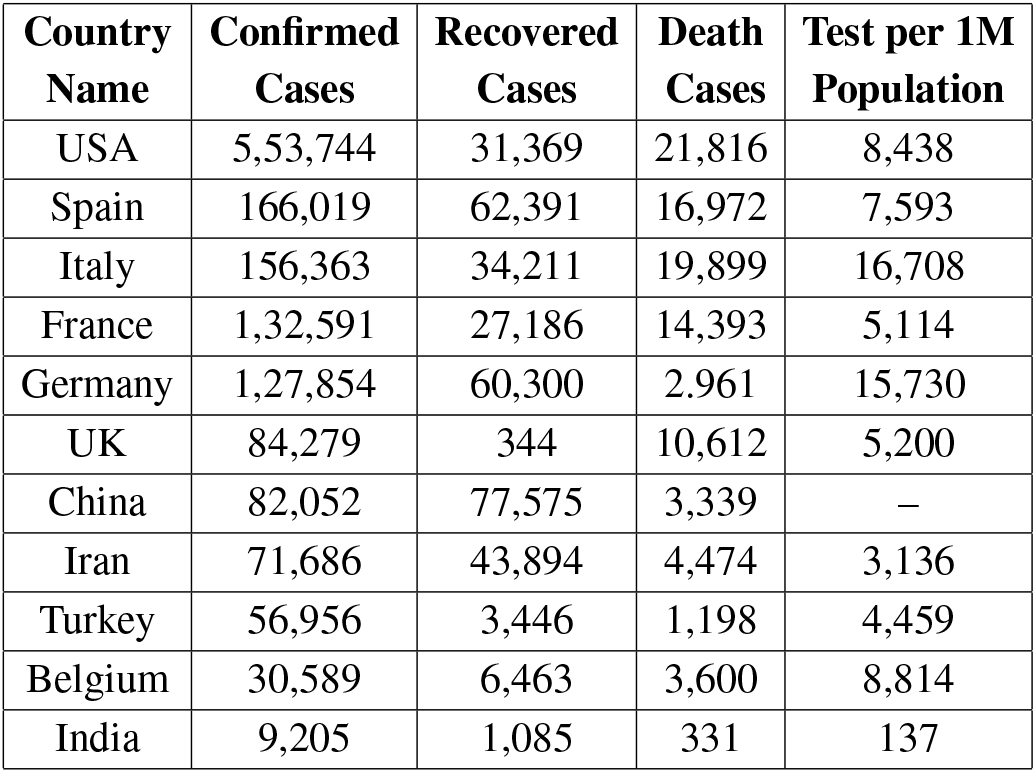
Country-wise (Top 10 & INDIA) statistical COVID-19 cases as of April 12, 2020^7^

| Country Name | Confirmed Cases | Recovered Cases | Death Cases | Test per 1M Population |
| --- | --- | --- | --- | --- |
| USA | 5,53,744 | 31,369 | 21,816 | 8,438 |
| Spain | 166,019 | 62,391 | 16,972 | 7,593 |
| Italy | 156,363 | 34,211 | 19,899 | 16,708 |
| France | 1,32,591 | 27,186 | 14,393 | 5,114 |
| Germany | 1,27,854 | 60,300 | 2,961 | 15,730 |
| UK | 84,279 | 344 | 10,612 | 5,200 |
| China | 82,052 | 77,575 | 3,339 | – |
| Iran | 71,686 | 43,894 | 4,474 | 3,136 |
| Turkey | 56,956 | 3,446 | 1,198 | 4,459 |
| Belgium | 30,589 | 6,463 | 3,600 | 8,814 |
| India | 9,205 | 1,085 | 331 | 137 |

**Figure 1.**
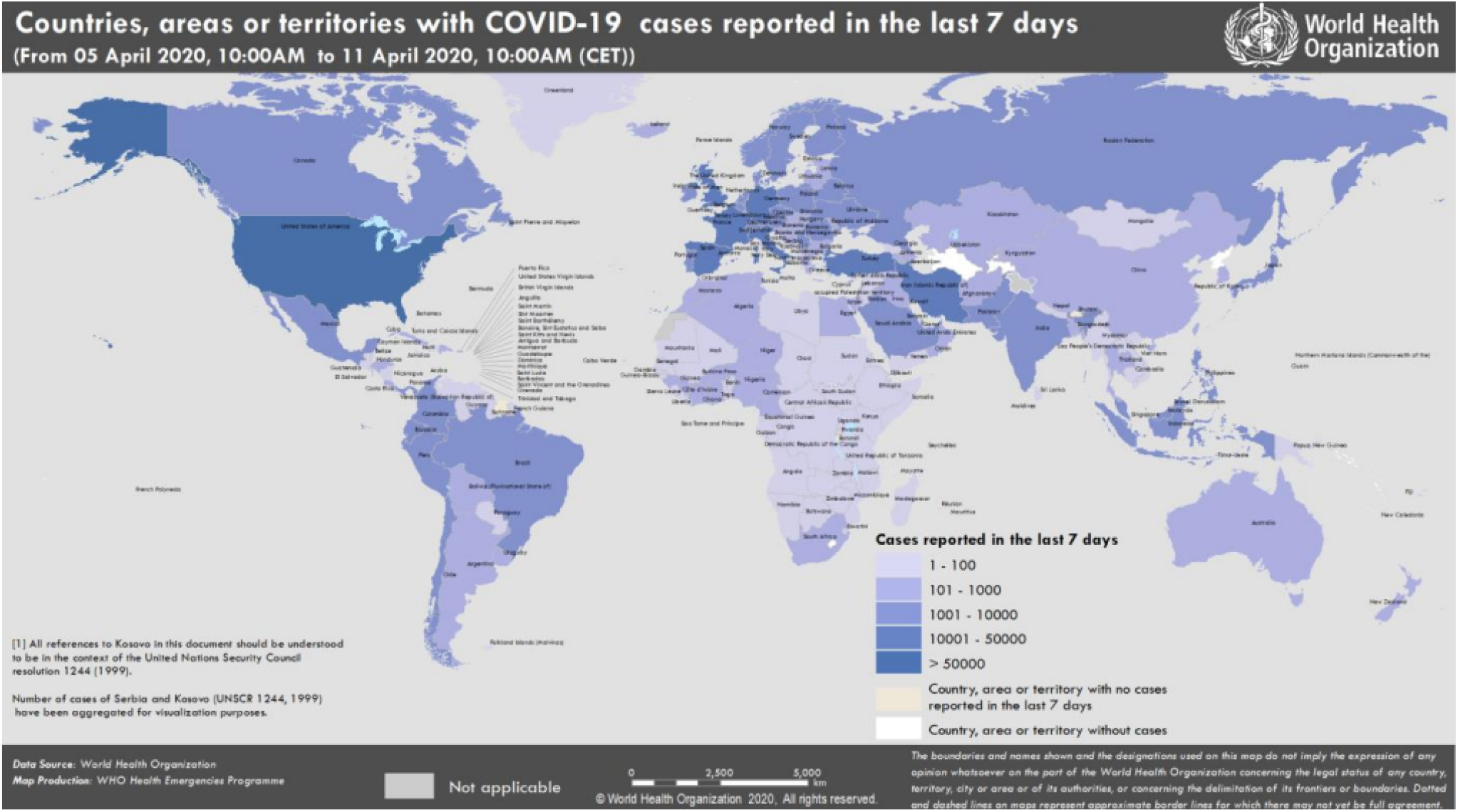
Countries, territories or areas with reported confirmed cases of COVID-19 as on April 12, 2020^8^.

Speaking of India, the country currently stands at 9, 205 confirmed cases and 331 deaths. There has been a 21-day complete lockdown in India as a measure to prevent further transmission, but whether India is coping up with the disease or not continues to be a big question. While these small numbers of confirmed and deceased cases may bring in a sigh of relief, they also challenge the state of medical testing and diagnosis in India^11^. Whether these numbers are low or they are low because of insufficient testing remains unanswered^12^.

In these times of emergency, researchers and scientists have come a long way in helping the society by indulging in scientific methods of identifying the virus inside humans through the technology of machine learning (ML) and deep learning(DL) algorithms.

In recently, several classical image processing and machine or deep learning methods are used to automatically classify the diseases with digitized chest X-ray images^13, 14^. Class decomposition of the coronavirus disease-2019 (COVID-19) in non-COVID and COVID viral infection with X-ray scans regarded as one of the critical subjects of matter for diagnosing this highly infectious disease^15–17^. For *eg*., One of the chest X-ray slides pictorially represented as COVID-19, Regular, and Pneumonia patient in Figure 4.

Fast COVID-19 recognition can help control disease transmission and will help to monitor the progression of infectious disease. According to Tao *et al*.^18^, it seems that Chest CT is more prone to COVID-19 diagnosis with respect to the original reverse-transcription polymerase chain reaction (RT-PCR) that has been collected from swab samples from patients and reported an accuracy of 97.3% to classify COVID-19 viral highly infectious diseases. Convolution neural network (CNN) is one of the most popular algorithms that have shown high precision in the ability to interpret the COVID-19 classification with medical images like X-rays or CT images. Wang *et al*.^19^ has proposed a COVID-19 classification technique by implementing the CNNs based on Inception Net over the pathogen-confirmed COVID-19, with 453 computed tomography (CT) images and reported an accuracy of 82.9%. Song*et al*^20^ has implemented a multi-class classification to recognise the diseases (COVID-19 viral infection, non-COVID and bacterial pneumonia) with CT images (88 patients with COVID-19 infected, 86 non-COVID patients and 100 patients with bacterial pneumonia)by using a modified version of pre-trained ResNet-50 net-work, named as DRE-Net and reported an accuracy of 86% for bacterial pneumonia and viral pneumonia (COVID-19) classification.

In another study, Sethy *et al*.^21^ used chest x-ray images to recognize COVID viral infection, firstly they have extracted deep features using CNN based on pre-trained ImageNETand at the last layer SVM were used in order to classify it. In addition, Wang *et al*.^16^ presented multi-class classification, deep convolutional neural network architecture named as COVID-Net implemented over 16, 756 chest radiography images that have been scanned with 13,645 patient to classify the COVID-19 and non-COVID also with a safe and bacterial infected patient.

## Results and Discussion

In this section, the results for the proposed methodology is discussed.

### Study Population Characteristics

Table 2 depicts the results of the final classification metrics produced by the proposed methodology. As can be seen in the table, many potential classifiers are listed which have been utilized for the performance calculation of the classification task. It can be inferred that the metrics corresponding to the Random Forest (RF) classifier and the XGBoost (XGB) classifier outperformed from the rest indicating that they have a better understanding of the image features which were input to them.

**Table 2.**
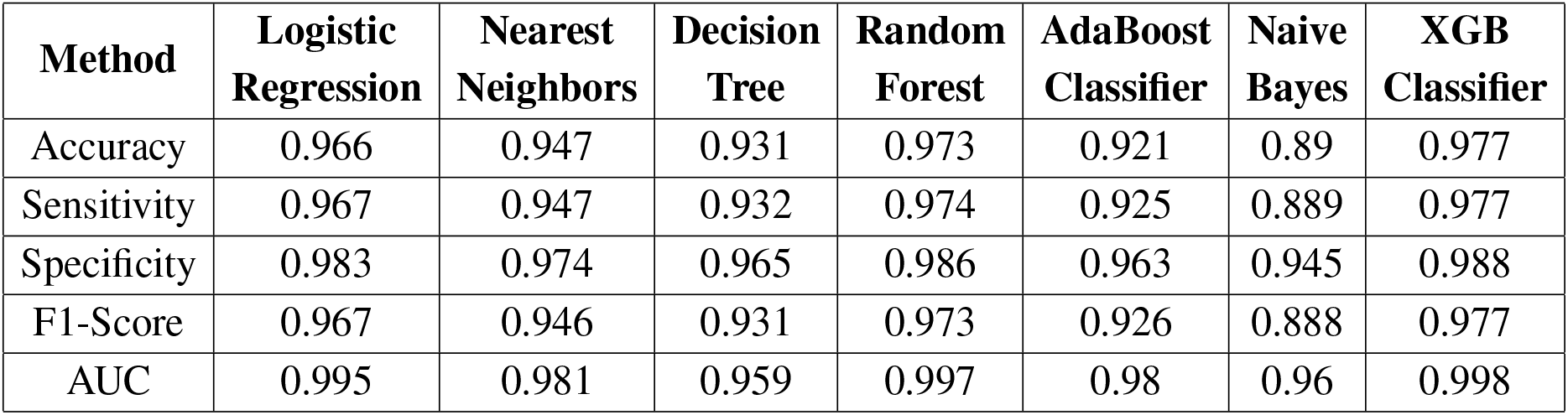
Performance Metrics of the Respective Classifiers

The corresponding performance metrics of RF and XGB in terms of Accuracy, Sensitivity, Specificity, F1-Score, and AUC are shown in Table 2. It seems that XGBoost performed the best among all the classifiers with an accuracy of 97.7%, while Logistic Regression (LR), k-Nearest Neighbour (kNN), Decision Tree (DT), Adaboost and Naive Bayes (NB) has an accuracy of 96.6%, 94.7%, 93.1%, 97.3%, 92.1% and 88.9%, respectively.

### Model Performance

Once the training of the model is performed, it becomes easy to further classify the features separately into 3 classes, namely, COVID-19, Pneumonia and Normal. In real-time, for example, if some random patient comes for screening, we can determine whether he/she has COVID or he/she is suffering from Pneumonia or Healthy using the proposed model by taking the chest X-ray and sending it to the proposed model.

To evaluate the efficiency of the proposed framework, confusion matrix is estimated, which gives a detailed understanding of the classification process in Figure 2. The classification model’s usefulness and productivity was measured using the traditional metrics of accuracy, precision, and recall. Precision is the calculation of the model’s correct predictions all over all predictions. Corresponding graphs of the Receiver Operating Characteristics (ROC) curves are being depicted as in Figure 3.

**Figure 2.**
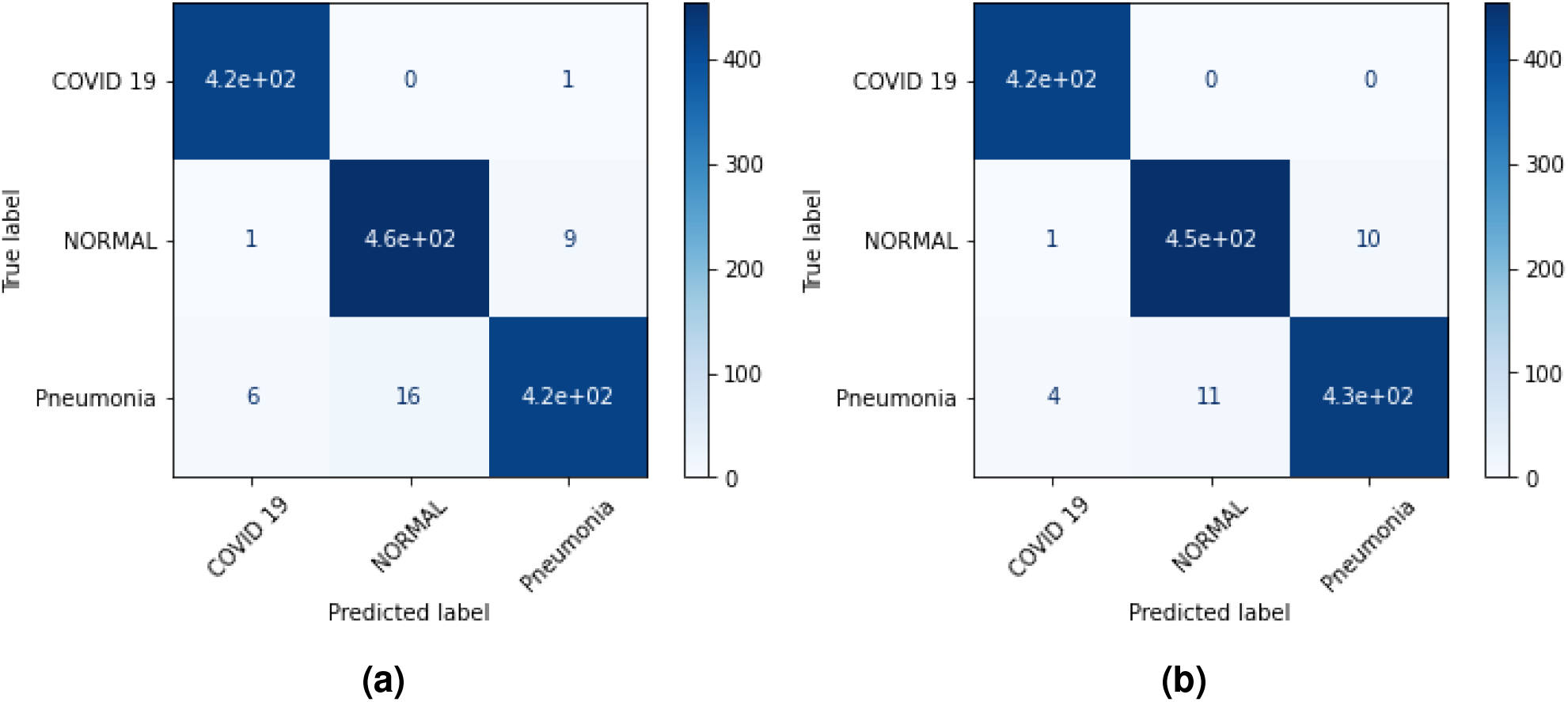
Confusion matrix of of the top two accuracy of the model obtained by respective classifiers (a) RF (b) XGB with respect to COVID-19, Normal, and Pneumonia patient chest x-ray images.

**Figure 3.**
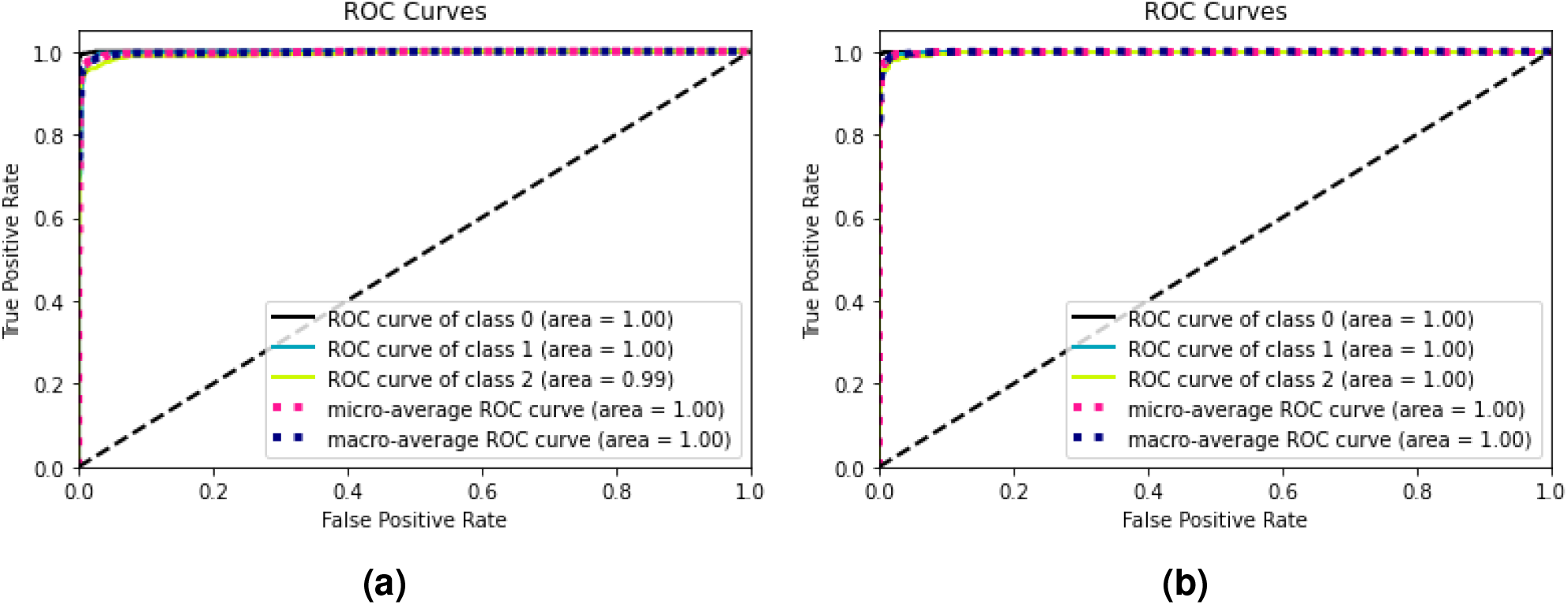
ROC-AUC curve of the top two accuracy of the model obtained by respective classifiers (a) RF XGB with respect to COVID-19, Normal, and Pneumonia patient chest x-ray images.

## Method

### Concoction of Dataset

We instantiated our proposed methodology with two publicly available datasets: Chest X-Ray Images (Pneumonia)^1^ and COVID-19 public dataset from Italy ^2^. For the training of the ResNet152 model architecture, a total of 5840 images are used with 5216 images for training (1341 for Normal class and 3875 for Pneumonia class) with the remaining 624 (234 for Normal class and 390 for Pneumonia class) in testing. For the final classification of COVID-19 patients by the machine learning classifiers, we used the images from the Chest X-ray Images (Pneumonia) dataset with 2748 images where 1833 (with 42 from COVID-19 patients, 894 in Normal patients and 897 in Pneumonia patients) images for training and 915 (with 20 from COVID-19 patients, 447 in Normal patients and 448 in Pneumonia patients) for testing whose samples are as shown in Figure 4. The number of the images used in the experiment from both the datasets are as depicted in Table 3 and Table 4.

**Figure 4.**
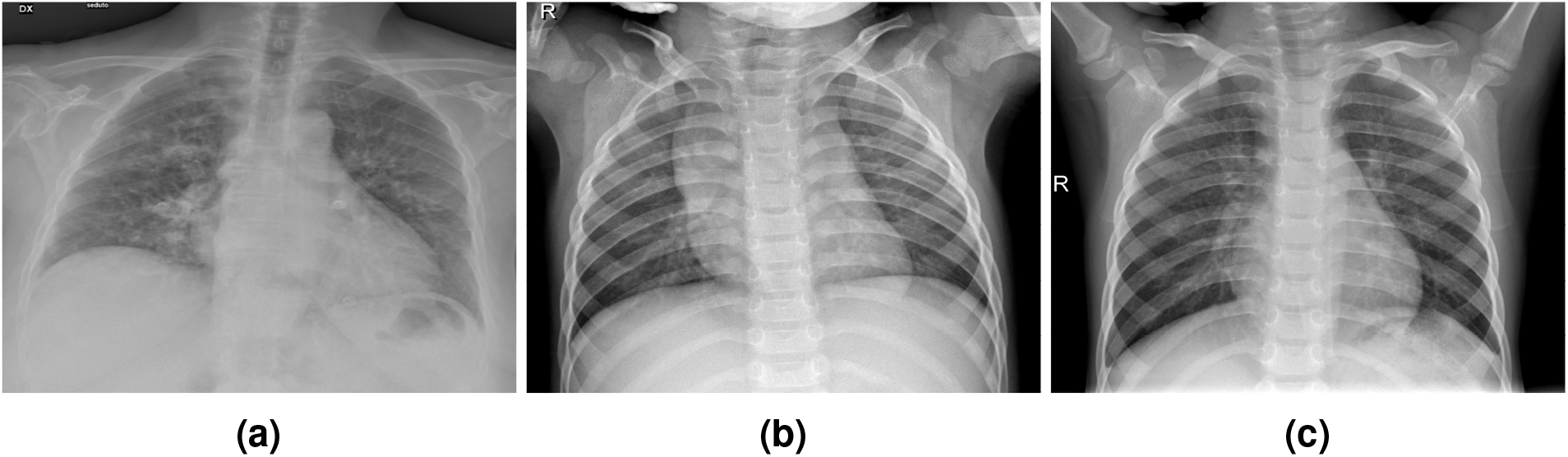
Examples of a) COVID-19 b) Normal, and c) Pneumonia, patient chest x-ray images.

**Table 3.** DATASET 1: Number of images for each class in COVID-19 dataset

| Dataset/Classes | COVID-19 | Normal | Pneumonia | Total |
| --- | --- | --- | --- | --- |
| # of Images in Training set | 42 | 894 | 897 | 1833 |
| # of Images in Test set | 20 | 447 | 448 | 915 |
| Total | 62 | 1341 | 1345 | 2748 |

**Table 4.** DATASET 2: Number of images for each class in Normal and Pneumonia patient dataset

| Dataset/Class | Normal | Pneumonia | Total |
| --- | --- | --- | --- |
| # of Images in Training set | 1341 | 3875 | 5216 |
| # of Images in Test set | 234 | 390 | 624 |
| Total | 1575 | 4265 | 5840 |

### Pre-processing of the Dataset

It could be seen that all the acquired X-ray images are of variable shapes and sizes, which increases the difficulty in effective classification. In order to effectively perform classification tasks, image pre-processing is performed. There exist many automatic and semi-automatic techniques for detecting abnormality in the body of the patient, but the absence of reliable and precise techniques can cause ambiguities in the classification process. Keeping aforementioned challenges in the mind, ML based predictive classifiers are used for analyzing the chest X-ray images, which are further discussed in the following sections.

X-ray images are taken with a low resolution which may have a variable height to width ratio. In order to facilitate training and testing of the deep networks, necessary pre-processing steps like image cropping and resizing is performed. In the proposed method, all input images are first converted to a standard size of 224 *×* 224 for a similar course of action in both the developed model.

### Architecture

The proposed framework for accurate prediction of COVID-19 using the chest X-ray images through deep feature learning model with SMOTE and machine learning classifiers consisted of the ResNet152 architecture for the training with later using the concerned features to classify the chest X-ray images using machine learning classifiers. The proposed framework is depicted in Figure 5. A detailed description of the architecture is explained below.

**Figure 5.**
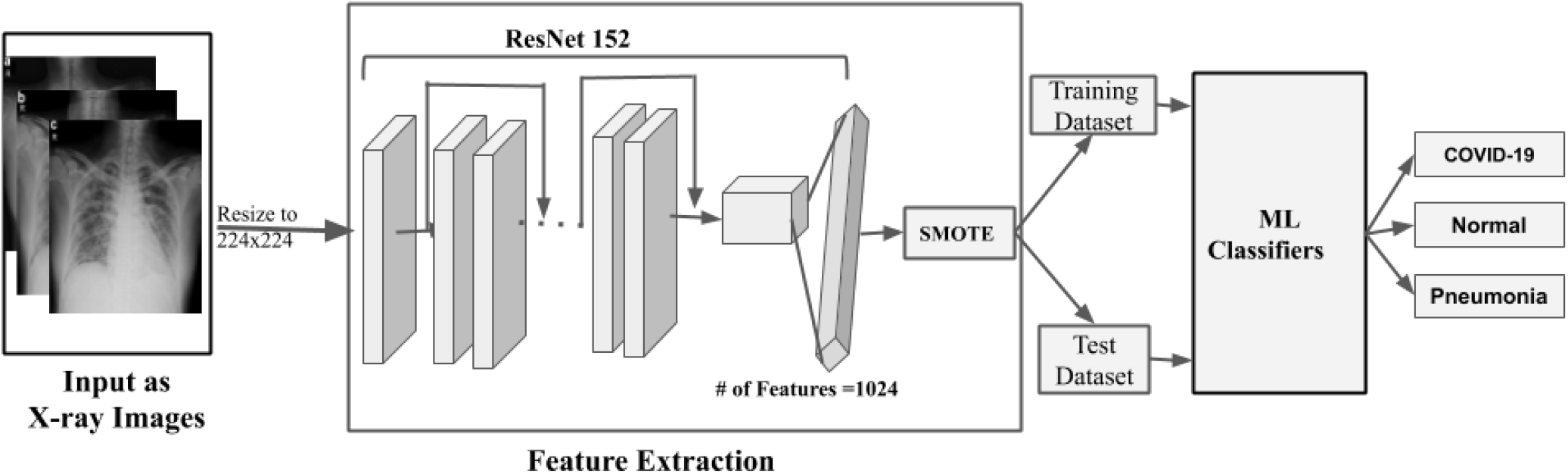
The block diagram of the proposed model for an early diagnosis of COVID-19

- The X-ray images of COVID-19, Normal and Pneumonia infected patients were taken and resized into the size of 224×224.
- A ResNet152 ^22^ model was trained for the classification of Pneumonia and Normal patients. ResNet is known to be a better deep learning architecture as it is relatively easy to optimize and can attain higher accuracy. Due to a large number of layers in the network architecture, it has high time complexity. This complexity can be reduced by utilizing a bottleneck design. Further, there is always a problem of vanishing gradient, which is resolved using the skip connections in the network.

Finally, the last fully connected (FC) layer of the network is followed by a Logarithmic Softmax layer with Adam optimizer to optimize the neural network.

- After completion of the training process, the last three layers from the ResNet152 were replaced with a ReLU, an FC layer, and an output layer. Now with the tweaked network, the last FC layer gives 1024 as output. The input images are now passed through this modified network to obtain 1024 features for each image in the dataset.
- Now, in the dataset, the number of data points for one class is very less when compared to the other class corresponding to the COVID-19 patients. Therefore, to balance the imbalanced data points, we use synthetic minority oversampling technique (SMOTE)^23, 24^. This algorithm creates an equal number of samples for each class. The above method was incorporated to ensure the smooth working of many machine learning algorithms like Logistic Regression, Decision Tree, etc. which otherwise tends to be more biased towards a majority class. The algorithm mentioned above generates virtual data points between existing points of the minority class by using linear interpolation.
- After processing all the images and converting them into features and using SMOTE for intra-class variations, the next step involves fitting the dataset using different machine learning predictive classifiers. For this purpose, We have integrated Logistic Regression (LR)^25^, k-Nearest Neighbour (kNN)^26^, Decision Trees (DT)^27^, Random Forest (RF)^28^, Adaptive Boosting (AdaBoost)^29^, Naive Bayes (NB)^30^ and XGBoost(XGB)^31^ to classify the COVID-19, Normal, and Pneumonia (shown in the Table 2).

### Experimental Setups

We have implemented the proposed classification system for COVID-19 diagnosis using Python 3.8 programming language with a processor of Intel^*®*^ Core i5-8300H CPU @ 2.30GHz *×*8 and RAM of 8 GB running on Windows 10 with NVIDIA Geforce GTX 1050 with 4GB Graphics.

### Evaluation criteria

To evaluate the efficacy of the model, the confusion matrix along with Area under Curve (AUC)^32^ are estimated and gives an understanding of the proposed methodology and its potential for detailed classification. The classification model’s usefulness and productivity were measured using the traditional metrics of accuracy, precision, and recall. Precision is the calculation of the model’s correct predictions all over all predictions. The classification of COVID-19 patients and Normal or Pneumonia patients and between Normal patients and Pneumonia is termed as Accuracy, Sensitivity, Specificity, and F1-score are represented mathematically in terms of confusion matrix as given in Equations 1, 2, 3, and 4, respectively.

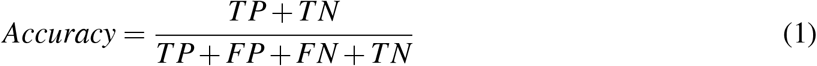

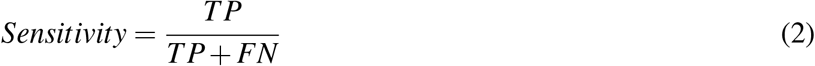

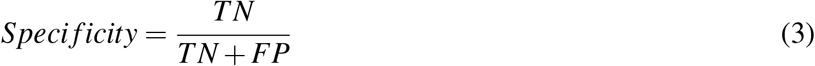

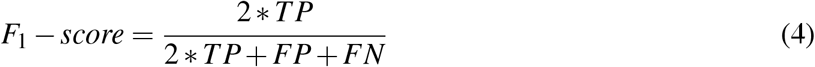

where TP, TN, FP, and FN are True Positive, True Negative, False Positive, and False Negative, respectively.

*1993CCZweig*

## CONCLUSION

In this work, we have presented the use of ResNet152 and machine learning classifiers for the effective classification of COVID-19. The proposed methodology is trained on two publicly available datasets and has outperformed across all the classes. We also encompassed the SMOTE algorithm for balancing the intra-class variation among the datasets. With the SMOTE based features, machine learning algorithms are applied for final classification leading to the best result obtained by Random Forest with the Accuracy, Sensitivity, Specificity, F1-score and AUC of 0.973, 0.974, 0.986, 0.973, and 0.997 (for Random Forest) and 0.977, 0.977, 0.988, 0.977, and 0.998 (for XGBoost), respectively. Therefore, this approach of using X-ray images and computer-aided diagnosis can be used as a massive, faster and cost-effective way of screening. Also, it brings down the time for testing drastically. To make a clinically effective prediction of COVID-19, training with more massive datasets and testing in the field with a larger cohort can be immensely useful.

## Data Availability

Dataset 1: Chest X-Ray Images (Pneumonia)
Dataset 1 Link: https://www.kaggle.com/paultimothymooney/chest-xray-pneumonia
Dataset 2: COVID-19 public dataset from Italy
Dataset 2 Link: https://towardsdatascience.com/covid19-public-dataset-on-gcp-nlp-knowledge-graph-193e628fa5cb
We are in the process of getting more data from confidential government sources.

## Acknowledgements

We acknowledge Consulate General of India, Osaka-Kobe for constant support and encouragement for this Bilateral India-Japan Artificial Intelligence (INJA-AI) module between the researchers of Indian Institute of Technology Roorkee (IITR), India and Kyoto University, Japan to better manage COVID-19 by non-invasive prediction.

## Footnotes

1 ^1^https://www.kaggle.com/paultimothymooney/chest-xray-pneumonia

2 ^2^https://towardsdatascience.com/covid19-public-dataset-on-gcp-nlp-knowledge-graph-193e628fa5cb

